# Evaluation of resistance of human head lice to pyrethroid insecticides: a meta-analysis study

**DOI:** 10.1101/2022.08.04.22278398

**Authors:** Ebrahim Abbasi, Salman Daliri, Zahra Yazdani, Shokrollah Mohseni, Ghulamraza Mohammadyan, Seyedeh Niloofar Seyed Hosseini, Reza Nasiri Haghighi

## Abstract

**Introduction:** Pediculosis is one of the most common annoying infections caused by insects in humans. Pyrethroids are one of the main insecticides used to treat this infection. But recently, due to the resistance of lice to this group of insecticides, its therapeutic effects have been affected. Based on this, the present study was conducted to investigating the prevalence of resistance to treatment against these insecticides in the world in the form of a meta-analysis.

**Methods:** This study was conducted as a meta-analysis on the prevalence of treatment resistance in human head lice against pyrethroid insecticides in the world. Based on this, all articles published without a time limit until the end of June 2022 in PubMed/MEDLINE, Web of Science (ISI), Scopus, and Google Scholar databases were extracted and using random effects model statistical methods in the meta-analysis, Cochrane, Index I2 and funnel plot were analyzed by STATA software.

**Results:** Twenty studies were included in the meta-analysis process. According to this the prevalence of resistance to pyrethroid insecticides in human head lice was estimated at 59% (CI95%: 50% to 68%). Among pyrethroid insecticides, the highest prevalence of resistance to treatment against permethrin insecticide was 65%. Regarding the prevalence of resistance by year, the prevalence before 2004 was estimated at 33%, but after 2015, this rate reached 82%. Also, the prevalence of resistance to treatment was estimated at 68% using genetic diagnosis methods and 43% using clinical diagnosis methods.

**Conclusion:** More than half of human head lice are resistant to pyrethroid insecticides. Based on this, it is recommended that before using this treatment method to treat human head lice infestation, the prevalence of resistance to treatment in that area should be investigated and if the prevalence of resistance is high, alternative or combined treatment methods should be used.

## Introduction

Pediculus humanus capitis or human head louse is an Infestation caused by a hematophagous ectoparasite that lives in human hair and feeds on human blood. Infestation with this ectoparasite is often asymptomatic and not pathogenic for humans, but it causes skin irritation and sometimes leads to skin infection due to skin itching (1, 2). But the importance of this pollution is its widespread prevalence, so that in recent decades, most of the countries of the world in North America, South America, Australia, Europe, and Asia have been involved in this pollution, and its prevalence is increasing (3-6). As a result, controlling and preventing the spread of this Infestation is very important. The transmission of this Infestation is from person to person, so in case of contact with an infected person, especially in crowded places, this Infestation is transmitted to other people and causes them to become infected (7, 8). One of the most important approaches to the prevention and control of Pediculosis is the treatment of infected people. Because when the source of contamination is controlled, the disease will no longer be transmitted. One of the most important and common treatment methods is the use of safe insecticides. Today, various insecticides are used in the world to treat Pediculosis, but the most common group of insecticides are Pyrethroids, which are sold and used under the trade names Permethrin, Phenothrin, and Pyrethrin (9-11). This group of insecticides causes their death by disrupting the nervous system of lice, and in addition to treating patients; it also interrupts the cycle of disease transmission. But recently, due to indiscriminate use and incomplete treatment, lice have become resistant to this group of insecticides, and due to the lack of complete treatment of the source of Infestation, its prevalence is increasing (12-14). Studies have shown that the effectiveness of permethrin for the treatment of Pediculosis has decreased from 97% and phenothrin from 75% to less than 15%. This shows that the use of this group of insecticides is not effective (10, 11, 15). In lice, there are different mechanisms for resistance to treatment against insecticides, the main mechanism, especially against pyrethroid insecticides, is resistance at the target site, which is created by point mutations. This makes the real targets of an insecticide less sensitive to the active ingredient (16).

This mutation in the voltage-sensitive sodium channel gene (Vssc) leads to resistance in sodium channels (17-19). By acting on sodium channels, pyrethroid insecticides interfere with the opening and closing of these channels, and this action leads to paralysis and eventually the death of lice, But when these mutations occur, the sensitivity of lice to insecticides is reduced and the treatment with them will be incomplete or ineffective (13, 19, 20). As a result, it is very important to be aware of the prevalence of resistance to insecticides to adopt the appropriate treatment. There are various methods to detect the prevalence of resistance in lice. The simplest method is to use insecticides to kill live lice and finally observe the proportion of live lice (clinical method). The next method is to use molecular indicators to determine the presence of treatment-resistant genes in lice (genetically method). In this method, after extracting the genes in the laboratory environment, the existence of gene mutation of resistance to the treatment and its ratio in lice are determined, this includes Polymerase Chain Reaction (PCR), Quantitative Sequencing (QS), and Serial Invasive. Signal Amplification Reaction (SISAR) Protocol (17, 21).

In general, it is necessary to be aware of the prevalence of treatment resistance in lice against pyrethroid insecticides to adopt the appropriate treatment. Based on this, the present study was conducted to determining the prevalence of treatment resistance in human head lice against pyrethroid insecticides via systematic review and meta-analysis in the world.

## Material and Methods

### Protocol

This systematic review and meta-analysis study was conducted by the Preferred Reporting Items for Systematic Reviews and Meta-Analyses (PRISMA) statement (22). This study aimed to estimate the prevalence of resistance to human head lice treatment against pyrethroid insecticides.

### Search strategy and study selection

The search for articles on the prevalence of resistance to the treatment of human head lice against pyrethroid insecticides was conducted in the scientific databases PubMed/MEDLINE, Web of Science (ISI), Scopus, and Google Scholar without time limit until the end of June 2022. to find the articles, the collection of related keywords extracted from medical subject headings (MeSH) including head lice, human head lice, Pediculosis, Pediculosis capitis, Human Head Louse, Insecticide, Pediculicide, resistant, Pyrethroids, pyrethrins, permethrin, phenothrin, Treatment was used. These keywords were searched using AND and OR operators in singular and compound form in titles, abstracts, and keywords. The reference lists of related articles were also checked manually for other possibly relevant articles that were not found through the electronic search strategy. Searching and extraction of articles were done independently by two trained authors in the field of searching articles in the medical information database.

### Inclusion and Exclusion criteria

All English language articles published in the field of human head louse resistance to treatment, which investigated the resistance to pyrethroid insecticides and the ratio of resistance reported in them and were of good quality, were included in the study. Articles with low quality, studies conducted on other insects except for human head lice, non-reporting of quantitative resistance to treatment, reviews of other insecticides, qualitative studies, case reports, case series, reviews, and meta-analysis were excluded from the study.

### Quality assessment

The Strobe checklist (Strengthening the Reporting of Observational Studies in Epidemiology) was used to evaluate the quality of the articles (23). This checklist has 22 sections in the fields of title, abstract, introduction, objectives, study method, Inclusion and exclusion criteria, sample size, study group, results, and limitations, which are scored based on the importance of each section. Be. The minimum and maximum scores that can be obtained by this checklist are 15 and 33. In this study, the minimum acceptable score was 20 (24).

### Data extraction

Two authors independently extracted the required information mentioned below from the articles: name of the author, year of study, place of study, sample size, type of insecticide used for treatment, method of detecting resistance, and prevalence of resistance.

According to the PRISMA flow chart (Figure 1), a total of 3451 studies were extracted during the search in scientific databases. Out of this number, 824 studies were removed due to repetition and from the remaining 2,627 articles, after reviewing the title and abstract, 2,554 articles were removed due to lack of relevance. Then the full text of the remaining 72 articles was studied. Of these, 52 articles were excluded for the following reasons: lack of investigation of pyrethroid insecticides (n=34), failure to report the prevalence of treatment resistance (n=15), and use of combined insecticides (n=3). Finally, 20 studies met the eligibility criteria and entered the systematic review and meta-analysis process.

**Figure 1.**
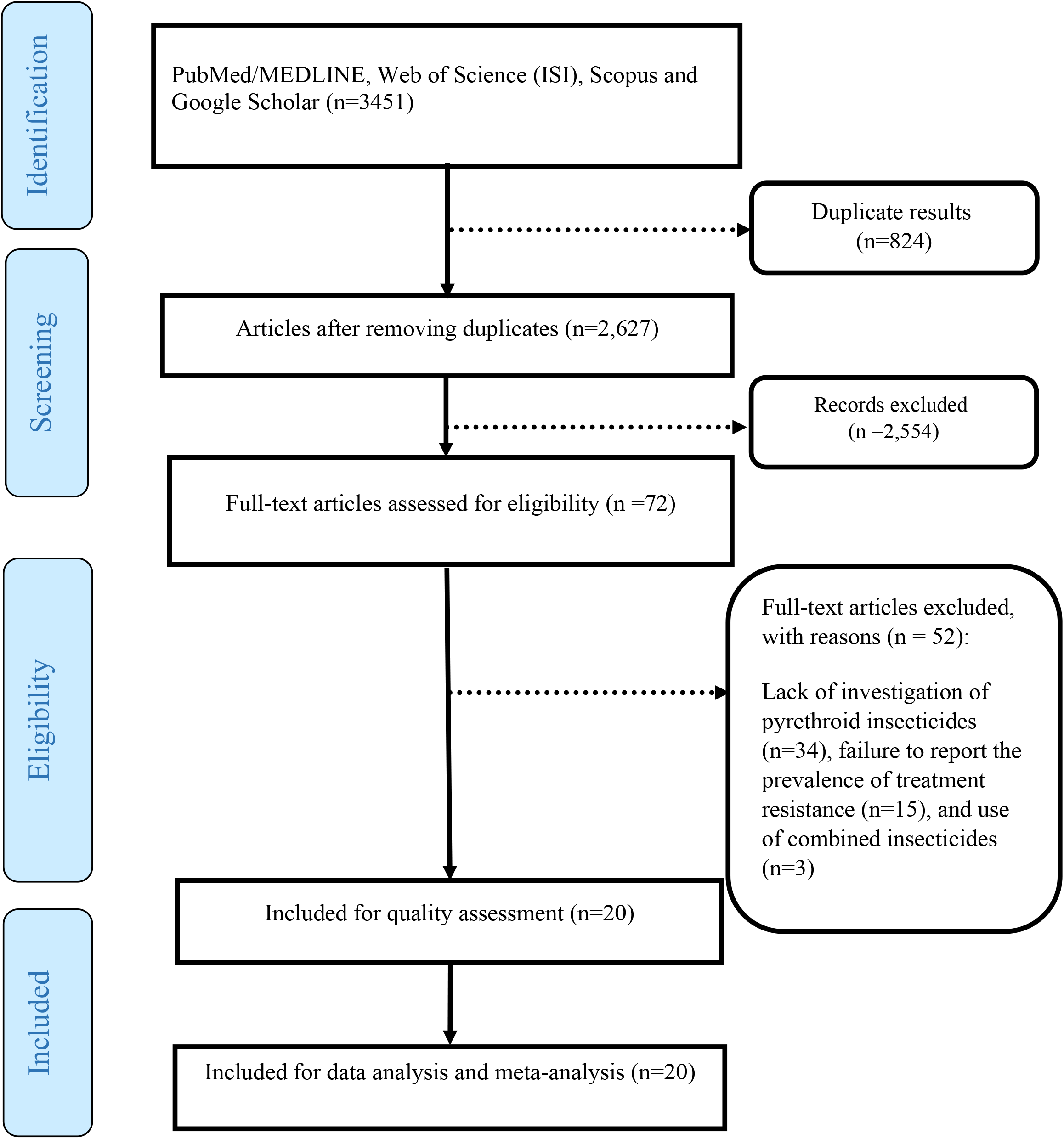
The PRISMA flow diagram

### Statistical analysis

Random effects model with 95% confidence interval was used to compare the prevalence of resistance. Analysis of subgroups was done based on the type of treatment, diagnosis, method and year of study. To evaluate the heterogeneity between the selected studies, the I2 test was used. In the results, 0%-25% heterogeneity was considered as low, 25%-50% as a medium, and 50%-75% as high heterogeneity. Publication bias was assessed by Begg-Mazumdar and Egger tests. In addition, sensitivity analysis was performed to assess the stability of the results. P-values above 0.05 indicate that the total variance is due to within-study variance rather than between-study variance. Stata V.14 (Stata Corp.) was used for statistical analysis.

## Result

Twenty studies conducted between the years 1991 and 2020 with a sample size of 24,651 human head lice were included in the meta-analysis process. The highest prevalence of human head lice resistance against pyrethroid insecticides was 99% and the lowest prevalence was 5.4%. The characteristic of the reviewed articles is presented in Table 1.

**Table 1.** characteristic of the articles included in the systematic review.

| Author | Place of study | Year of study | Sample size | Prevalence<br>Resistance | Quality assessment |
| --- | --- | --- | --- | --- | --- |
| Mumcuoglu KY (25) | Israel | 1991 | 50 | 21.2 | Mild |
| Thomas D (26) | United Kingdom | 2006 | 316 | 5.4 | High |
| Burgess IF (11) | United Kingdom | 2013 | 47 | 85.1 | Mild |
| Kim HJ (21) | USA | 2004 | 27 | 56 | Mild |
| Kasai SH (27) | Japan | 2009 | 630 | 9 | High |
| Yoon KS (28) | Netherlands and<br>United States | 2014 | 33 | 99 | High |
| Bouvresse S (29) | France | 2012 | 670 | 98 | High |
| Kasai SH (30) | Japan | 2009 | 282 | 9 | Mild |
| Eremeeva ME (14) | USA | 2017 | 259 | 99 | High |
| Yoon KS (31) | North American | 2014 | 480 | 99 | High |
| Audino G (32) | Argentina | 2005 | 240 | 36 | Mild |
| Gao JR (33) | USA | 2003 | 339 | 41 | High |
| Firoozian S (19) | Iran | 2017 | 2765 | 59 | High |
| Durand R (34) | France | 2011 | 1551 | 93 | High |
| Tolozza AC (35) | Argentina | 2014 | 154 | 88 | High |
| Roca-Acevedo G (36) | Chile | 2019 | 99 | 49 | Mild |
| Ponce-Garcia G (37) | Mexico | 2017 | 468 | 62 | High |
| Gellatly KJ (38) | USA | 2016 | 14281 | 98 | High |
| Karakuş M (39) | Turkey | 2020 | 1767 | 99 | High |
| Chosidow O (40) | New Zealand | 1994 | 193 | 60 | Mild |

| Author | Place of study | Year of study | Sample size | Prevalence Resistance | Quality assessment |
| --- | --- | --- | --- | --- | --- |
| Mumcuoglu KY (25) | Israel | 1991 | 50 | 21.2 | Mild |
| Thomas D (26) | United Kingdom | 2006 | 316 | 5.4 | High |
| Burgess IF (11) | United Kingdom | 2013 | 47 | 85.1 | Mild |
| Kim HJ (21) | USA | 2004 | 27 | 56 | Mild |
| Kasai SH (27) | Japan | 2009 | 630 | 9 | High |
| Yoon KS (28) | Netherlands and United States | 2014 | 33 | 99 | High |
| Bouvresse S (29) | France | 2012 | 670 | 98 | High |
| Kasai SH (30) | Japan | 2009 | 282 | 9 | Mild |
| Eremeeva ME (14) | USA | 2017 | 259 | 99 | High |
| Yoon KS (31) | North American | 2014 | 480 | 99 | High |
| Audino G (32) | Argentina | 2005 | 240 | 36 | Mild |
| Gao JR (33) | USA | 2003 | 339 | 41 | High |
| Firoozian S (19) | Iran | 2017 | 2765 | 59 | High |
| Durand R (34) | France | 2011 | 1551 | 93 | High |
| Tolozza AC (35) | Argentina | 2014 | 154 | 88 | High |
| Roca-Acevedo G (36) | Chile | 2019 | 99 | 49 | Mild |
| Ponce-Garcia G (37) | Mexico | 2017 | 468 | 62 | High |
| Gellatly KJ (38) | USA | 2016 | 14281 | 98 | High |
| Karakuş M (39) | Turkey | 2020 | 1767 | 99 | High |
| Chosidow O (40) | New Zealand | 1994 | 193 | 60 | Mild |

Based on the findings of the meta-analysis of 20 studies, the prevalence of resistance to pyrethroid insecticides in human head lice was estimated at 59% (CI95%: 50% to 68%) (Figure 2). Among pyrethroid insecticides, the highest prevalence of resistance to treatment against permethrin insecticide was 65% (Figure 3).

**Figure 2:**
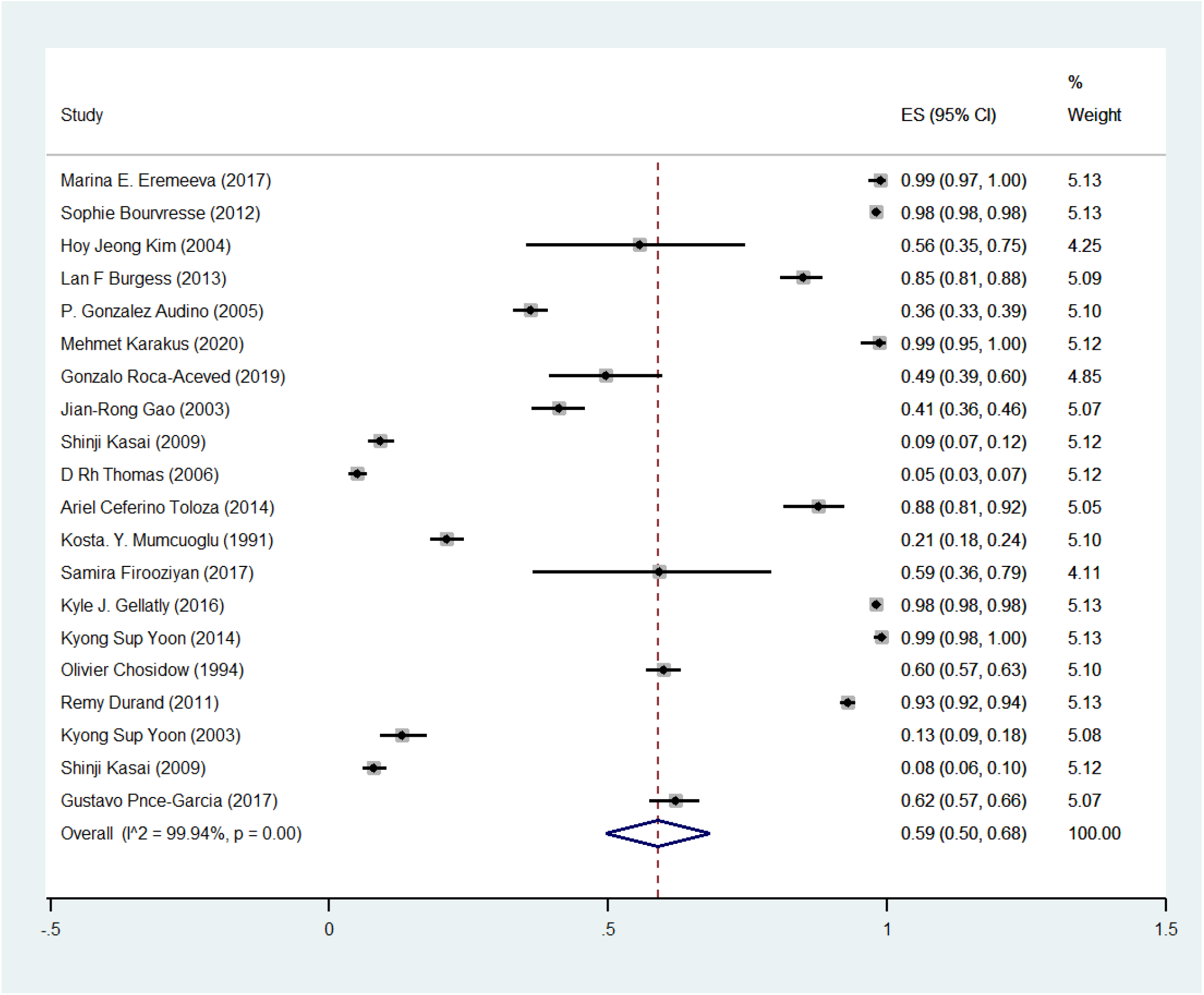
Pooled prevalence rate of Resistance to the treatment of human head lice against pyrethroid insecticides based on random effects model. The midpoint of each line segment shows the prevalence estimate, the length of the line segment indicates the 95% confidence interval in each study, and the diamond mark illustrates the pooled prevalence of resistance.

**Figure 3:**
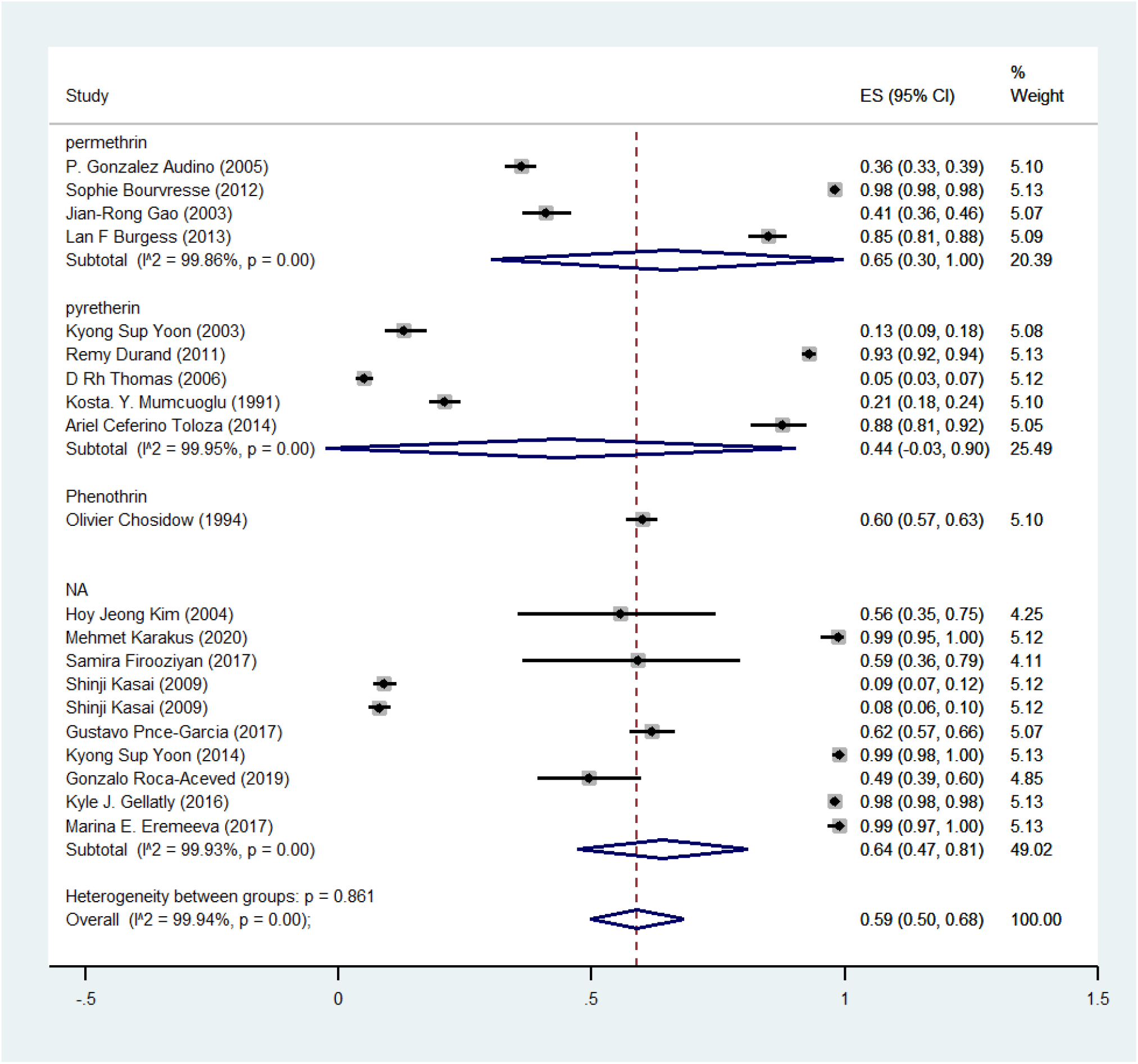
Pooled prevalence rate of Resistance to treating human head lice based on the type of insecticide using a random-effects model.

In the field of the method of detecting resistance to treatment in human head lice, molecular methods were used in the studies and the death rate of head lice after treatment was used. Based on this, the results of the meta-analysis showed that in the studies that used the genetic diagnostic method, the prevalence of resistance to treatment was 68%, and in the studies that used the clinical method (rate lice live after use insecticide), the prevalence of resistance was 43%, and in the studies that used both methods, the prevalence of resistance to treatment was estimated at 49% (Figure 4). Also, regarding the prevalence of resistance to treatment based on the year of the study, the studies were divided into three 10-year periods including before 2004, 2005 to 2015, and after 2015. Based on the findings of the meta-analysis of studies, the prevalence of resistance to treatment was estimated at 33% before 2004, 68% between 2005 and 2015, and 82% after 2015 (Figure 5).

**Figure 4:**
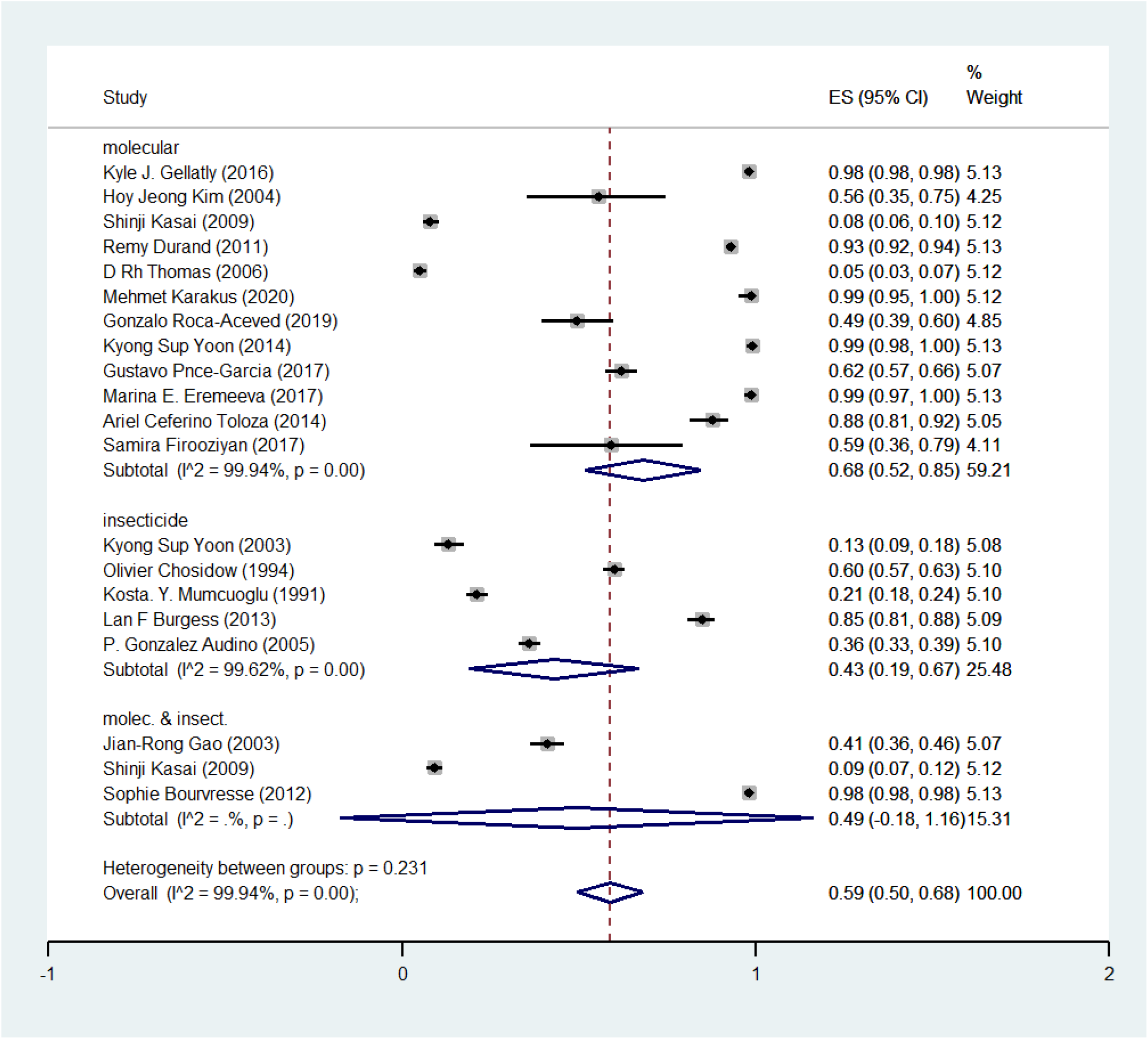
Pooled prevalence rate of Resistance to treating human head lice based on the Method for diagnosing resistance using a random-effects model.

**Figure 5:**
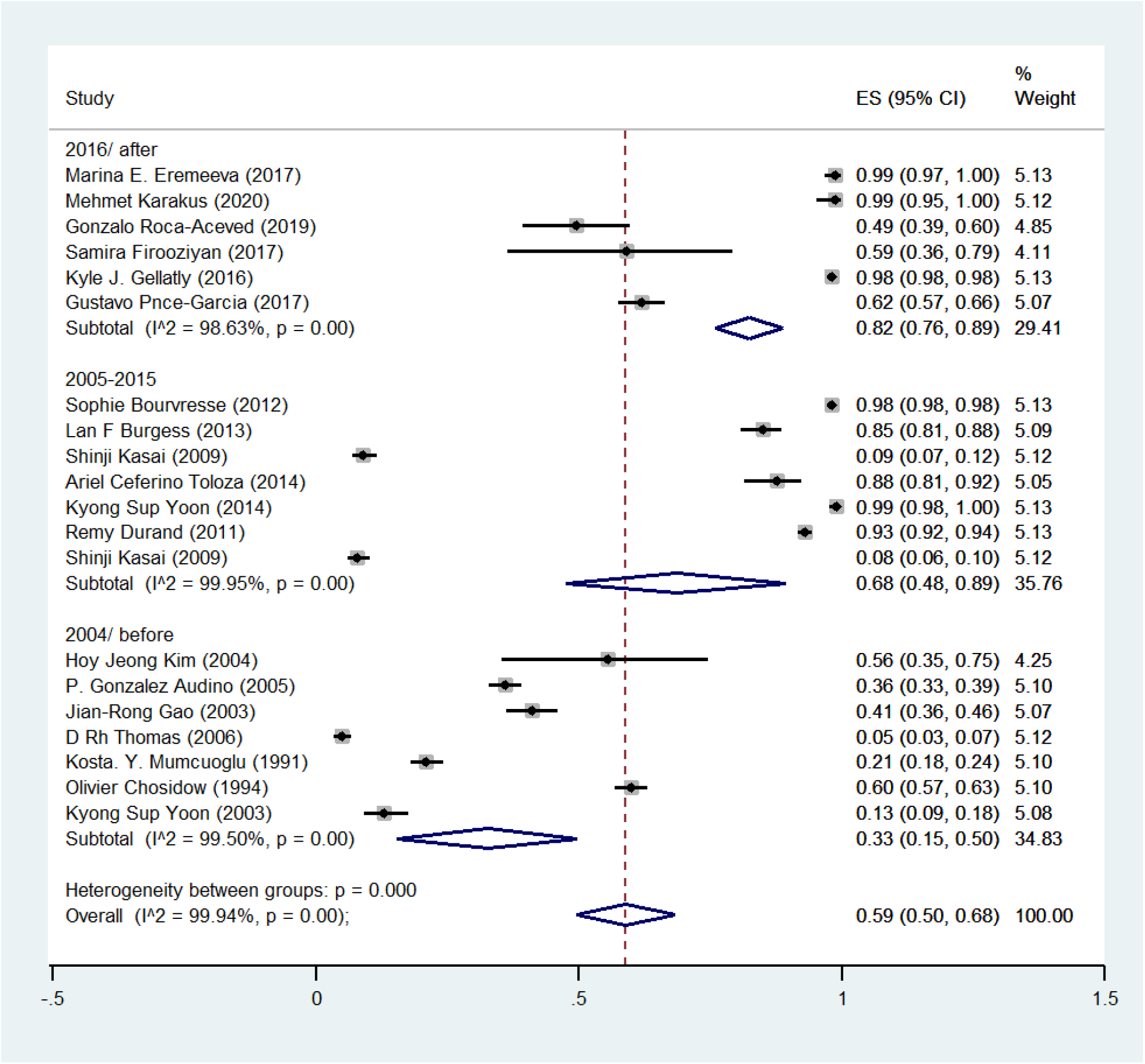
Pooled prevalence rate of resistance to human head lice treatment based on the year of the study using the random-effects model.

Funnel plot, Begg’s, and Egger’s tests were used to investigate the Publication bias. The symmetry of the articles in the funnel plot indicates that publication bias has not occurred (Figure 6). Also, the results of Begg’s and Egger’s tests show of also showed that there is no publication bias in this meta-analysis. The result of Begg’s test was Z=-0.39 and P=0.75 and the result of Egger’s test was Z=1.60 and P=0.11.

**Figure 6.**
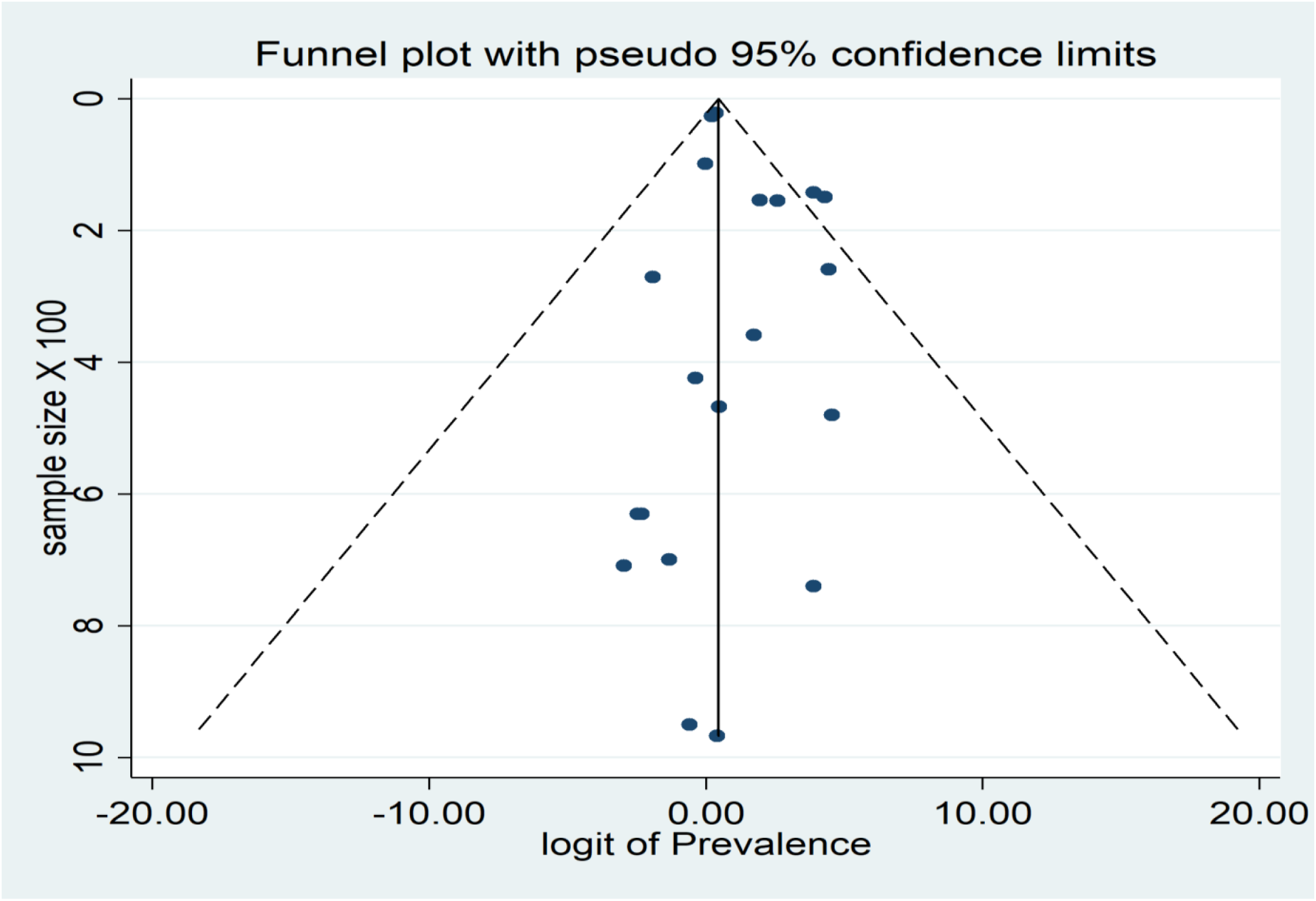
Funnel plot of the prevalence of of human head lice resistance

## Discussion

The findings of the present study showed that more than half of human head lice are resistant to pyrethroid insecticides, and the highest prevalence of resistance to permethrin insecticide was observed. Also, the prevalence of resistance to treatment had increased during the past three decades. In the study of Larkin et al (2020) (41) in Honduras 93.9%, in the study of Brownell et al (2020) (42) in Thailand 40%, and the study of Roca Acevedo et al (2019) (36) in Chile 50% of head lice had a mutation in the T917I allele, which indicates resistance to the treatment against pyrethroid. Ponce et al. (2017) reported in Mexico that 78.2% of the samples had a mutation in the T929I allele, which leads to resistance to treatment against pyrethroid insecticides (37). In the study by Gellatly et al. (2016), 98.3% of the lice studied in the United States had an allele resistant to pyrethroid treatment (38). Yoon et al (2014) in North America investigated the trend of resistance for 10 years in North America. Based on the findings, between 1999 and 2009, the prevalence of resistance to the treatment of Pyrethroids was estimated at 84.4%. This proportion was reported as 97.1% in 2008 and 99.6% between 2007 and 2009, which indicates the increasing trend of treatment resistance (31). Kasai et al (2009) stated that a large population of head lice in Japan is resistant to pyrethroid (27). Cueto et al (2008) reported resistance to permethrin in embryos and lice eggs in Argentina (43). In the study of Kristensen et al (2006) in Denmark, the study of head lice showed that the genetic mutation of alleles T929I and L932 was present in them and caused resistance to permethrin and Malathion (44). Yoon et al (2004) observed resistance to permethrin and Malathion in head lice in California and Florida (45). Gao et al (2003) reported resistance to permethrin caused by T929I and L932F alleles in California, Texas, and Florida (33). Lee et al (2000) reported resistance to permethrin and Malathion in England (46). Durand et al (2014) in France, considering the high rate of head lice genotypic resistance against pyrethroid insecticides, did not find its use effective for the treatment of head lice and recommended the use of alternative treatment methods (34). Lebwohl et al. (2007) who investigated the treatment of Pediculosis, considering the 12-day period for head lice to mature, it was recommended that to use pediculicides that does not have ovulatory properties (such as Pyrethroids), the treatment should be carried out in 2 or 3 the period and treatment with pediculicide against which there is genetic resistance is ineffective (47).

The survival of head lice after the use of insecticides can have various reasons, including the patient’s non-adherence to the treatment protocol and incorrect treatment (low dose or incorrect use), lack of egg-laying properties, re-infection, and resistance against treatment. Excessive pressure in the field of widespread and incomplete use of insecticides to control lice is the main cause of increasing resistance to treatment in human head lice. Resistance to insecticides leads to an increase in the prevalence and chronicity of infection in people, followed by an increase in treatment costs, the ineffectiveness of insecticides, the use of additional treatments, and an increase in possible toxicity and discomfort for infested people (48). Based on this, choosing the right insecticide and the correct prescription and treatment is essential in controlling the spread of human head lice and reducing the prevalence of resistance to treatment.

Different methods are used to determine the proportion of resistance to treatment in lice, which include: 1-Diagnosis of clinical resistance, in the form of the proportion of surviving lice 1 day after the use of insecticide, 2-Diagnosis of parasitic resistance, in the form of evaluation of the resistance proportion of lice outside the human body to pediculicide compounds and 3-diagnosis of genetic resistance, evaluation in the form of evaluation of polymorphism in genes related to resistance in lice. The findings of the present study showed that the prevalence of resistance using genetic diagnostic methods was higher than the prevalence using the proportion of dead lice after using insecticides. One of the reasons for its decrease can be pointed to incorrect treatment such as low dose of insecticide or incorrect use of medicine and lack of ovulatory properties. For genetic diagnosis, Karakuş et al. (2020) use the polymerase chain reaction (PCR) method to screen the mutation of resistant alleles in lice, which requires small amounts of DNA for analysis and is easy to perform in a laboratory environment recommended (14, 18, 39). Lee et al recommended the Quantitative Sequencing (QS) method for screening the mutation of treatment-resistant alleles in head lice due to its speed, accuracy, simplicity, and widespread use (49). Kim et al in Argentina recommended the use of serial invasive signal amplification reaction (SISAR) protocol for screening and diagnosis of treatment resistance (21). In general, although genetic methods are more accurate for diagnosing resistance to treatment, clinical methods are also a suitable option due to their availability, ease and cheapness.

## Limitations

The limitations of the present study include 1-the uncertainty of the type of insecticide used in some studies, 2-conducting studies in different years and different countries, which may have different treatment approaches. 3-The existence of heterogeneity between studies.

## Conclusion

The findings of the present study showed that more than half of human head lice are resistant to treatment with pyrethroid insecticides. Based on this, widespread use without specific treatment resistance in infected areas is ineffective. Based on this, it is recommended that before using this treatment method to treat human head lice infestation, the prevalence of resistance to treatment in that area should be investigated and if the prevalence of resistance is high, alternative or combined treatment methods should be used.

## Data Availability

All relevant data are within the manuscript and its Supporting Information files.

https://www.crd.york.ac.uk/prospero/display_record.php?RecordID=215924

## Declaration

### Ethics approval and consent to participate

Not applicable.

### Availability of data and materials

All the data obtained from this study are included in the text of the article.

### Authors’ contributions

EA determined the title, wrote and registered the protocol, and submitted the article. EA and SM extracted the files from the databases. ZY, SH, and RN, screening, and selection of final articles. GM and Abbasi, data extraction. SD wrote the article. All authors read and approved the final manuscript.

## Acknowledgements

We are grateful to Dr. Abbas Keshtkar, a member of the academic staff of Tehran University of Medical Sciences, for his statistical and practical advice.

